# A Bayesian Perspective Extracorporeal CPR for Refractory Out-of-Hospital Cardiac Arrest

**DOI:** 10.1101/2023.02.13.23285890

**Authors:** James M Brophy

**Author notes:** Email address (James M Brophy MD PhD), JMB is a research scholar supported by Les Fonds de Recherche Québec Santé.

## Abstract

**Background:** Whether extracorporeal CPR (eCPR) has survival benefits over conventional CPR (cCPR) in patients with refractory out-of-hospital cardiac arrest is an unresolved clinical question. Performing trials in this environment is exceedingly challenging and inferences need careful examination.

**Objective:** Determine if a Bayesian perspective provides additional inferential insights.

**Methods:** The INCEPTION trial of patients with refractory out-of-hospital cardiac arrest reported eCPR and cCPR had similar effects on the primary outcome, 30 day survival with a favorable neurologic outcome. Herein the probability of eCPR superiority, equivalence or inferiority to cCPR is re-evaluated with a Bayesian analysis using both vague and informative priors (from previously completed randomized clinical trials (RCTs)).

**Results:** Depending on the chosen prior, the Bayesian reanalysis of the INCEPTION intention-to-treat (ITT) data suggests an equivalence probability < 10% (defined as an absolute risk difference (RD) < 1%) but a clinical superiority probability of 66 - 99 % (defined as RD > 1.0). An INCEPTION per protocol (PP) analysis with a vague prior suggested a 1% probability of clinical benefit but this posterior probability increased to 86% when informative PP data from previous RCTs were considered.

**Conclusion:** Bayesian INCEPTION trial re-analyses provide additional quantative insights. The totality of the ITT evidence reveals a high probability for a clinically meaningful eCPR benefit over cCPR at 30 days. A PP analysis shows a less definitive probability of benefit. (Abstract word count 197, Manuscript word count 1477)

## Introduction

Out-of-hospital cardiac arrest is a frequent event and its devastating consequences are partially mitigated by rapid commencement of basic life support with high-quality chest compressions and external defibrillation (conventional cardiopulmonary resuscitation (cCPR)). However, there remains a substantial subset of individuals who do not respond rapidly to these measures and whether the addition of extracorporeal CPR (eCPR) to standard cCPR can improve survival and diminish anoxic brain injury is a current topic of research. The INCEPTION randomized clinical trial (RCT) recently published their results(Suverein et al. 2023) and concluded “In patients with refractory out-of-hospital cardiac arrest, extracorporeal CPR and conventional CPR had similar effects on survival with a favorable neurological outcome”.(Suverein et al. 2023)

The goal of this communication is to examine whether a Bayesian perspective permits additional inferential insights regarding the added value of eCPR in patients refractory to cCPR.

## Methods

The data for the primary outcome, 30 day survival with intact neurological status, based on an intention to treat (ITT) analysis was abstracted from the original INCEPTION trial (Suverein et al. 2023) and used for the primary analysis. A per protocol (PP) analysis accounts for adherence by analyzing only those patients who completed the treatment they were originally allocated to and while providing insights into efficacy may be subject to bias. A secondary analysis with PP data, abstracted from INCEPTION Figure S4(Suverein et al. 2023), was also performed.

In a Bayesian analysis probability statements arise from Bayes Thereom, expressed as:

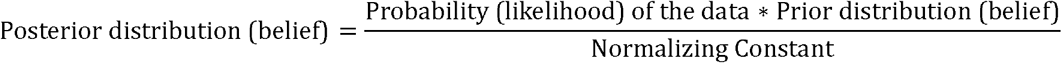

Therefore, in addition to the probability of the current data (likelihood function), prior probability distributions are required to form our posterior probability beliefs. Because our main focus is the analysis and interpretation of the INCEPTION trial(Suverein et al. 2023), our primary analysis used a default vague beta(*α, β*) distribution with a 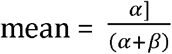 equal to the proportion of successes, and a 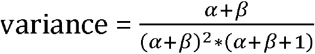 prior, beta(1,1). This assures that the posterior distribution is dominated by the observed INCEPTION(Suverein et al. 2023) data.

The robustness of the Bayesian approach can be assessed by considering different prior distributions. Incorporating prior information underscores another important advantage of Bayesian analyses, the ability to learn sequentially. There were two previous RCTs examining extracorporeal CPR(Yannopoulos et al. 2020; Belohlavek et al. 2022) and while the protocols are not completely identical, it is reasonable to allow this data to serve as informed priors, which can be updated with the INCEPTION(Suverein et al. 2023) data.

Therefore besides the vague prior, we considered three additional informative priors

1. an enthusiastic prior, so labelled since this uses only the ARREST(Yannopoulos et al. 2020) data, a trial stopped prematurely for efficacy
2. a skeptical prior, so labelled since this uses only the PRAGUE(Belohlavek et al. 2022) data, a trial stopped prematurely for futility for their primary 6 month outcomes
3. a combined prior using all the available prior RCT data(Yannopoulos et al. 2020; Belohlavek et al. 2022)

This prior probability of success in each arm (cCPR and eCPR) for each previous trial can also be summarized as a beta(*α, β*) distribution where *α* − 1 is the number of successes and *β* − 1 as the number of failures.

Posterior distributions are summarized with medians and 95% highest-density intervals (credible intervals (CrI)), defined as the narrowest interval containing 95% of the probability density function (McElreath 2020). Bayesian analyses permit not only calculations of the posterior probability of any additional survival with eCPR, but also of clinically meaningful benefits. While there is no universal definition for a clinically meaningful benefit, an absolute risk difference (RD) > 1% may be an acceptable threshold for many. Bayesian analyses also allows calculation of the probability between any two points. Consequently, we calculated a range of practical equivalence (ROPE) between treatments. While different ranges may be proposed, an absolute RD of +/- 1% seems a reasonable small difference that many would consider as equivalent.

Posterior distributions were estimated by fitting a binomial regression model with a single treatment parameter using the identity link, thereby allowing the calculation of risk differences between the treatments. Analyses were performed using the brms package(Bürkner 2017) within the integrated development environment of RStudio. Model convergence was assessed by examination of the Monte Carlo Standard Error < 10% of the posterior standard deviation, *n*_*eff*_ an estimate of the effective number of independent draws from the posterior distribution of the estimand > 10% maximum and 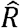 a measures the ratio of the average variance of samples within each chain to the variance of the pooled samples across chains < 1.1. Reporting followed the Bayesian Analysis Reporting Guidelines(Kruschke 2021). The statistical code can be found on Github (https://g.ithub.com/brophyj/eCPR).

## Results

A Bayesian analysis of the INCEPTION(Suverein et al. 2023) trial, which reported 10 and 14 successes among 62 cCPR and 70 eCPR patients respectively (p=0.52), was first performed with a default vague prior. This gives a risk difference (RD) of 3.8% (95% credible interval (CrI)-9.5 - 17.1) which can be transformed to an odds ratio (OR 1.3, 95% CrI 0.5 - 3.2). The closeness of this result to the original published analysis (OR 1.4, 95% CI 0.5 - 3.5) confirms the minimal impact of the default vague prior and reveals the Bayesian analysis is completely dominated by the observed INCEPTION(Suverein et al. 2023) data.

The eCPR probability density function for improved 30 day survival from INCEPTION(Suverein et al. 2023) data with the default vague prior is displayed graphically in Figure 1A and reveals that the probability of enhanced survival with eCPR is 71.2%. The probability that the improved survival exceeds a 1% RD improvement is 66% and the ROPE probability is 10% (Table 1).

**Table 1.** eCPR risk differences 95% credible intervals and probabilities with various priors ITT analyses.

| Priors | ITT analyses |  |  |  |  |  |
| --- | --- | --- | --- | --- | --- | --- |
|  | RD | 95% CrI |  | Probabilities |  |  |
|  | point estimate | lower limit | upper limit | p(RD) >0 | p(RD) >1% | p(ROPE) |
| Vague | 3.8 | -9.5 | 17.1 | 71.2 | 66.0 | 10.0 |
| Enthusiastic | 9.5 | -2.8 | 21.9 | 93.5 | 91.2 | 4.1 |
| Skeptical | 9.2 | 0.9 | 17.5 | 98.5 | 97.4 | 1.8 |
| Combined | 11.1 | 2.9 | 19.3 | 99.6 | 99.2 | 0.6 |

Vague: default vague prior
Combined: prior eCPR data from ARREST + PRAGUE
Enthusiastic: prior eCPR data from ARREST alone
Skeptical: prior eCPR data from PRAGUE alone
(PRAGUE trial stopped for futility for 6 month outcomes, but 30 day outcomes supportive of eCPR
ROPE: range of practical equivalence = + / - 1% RD (risk difference)

**Figure 1.**
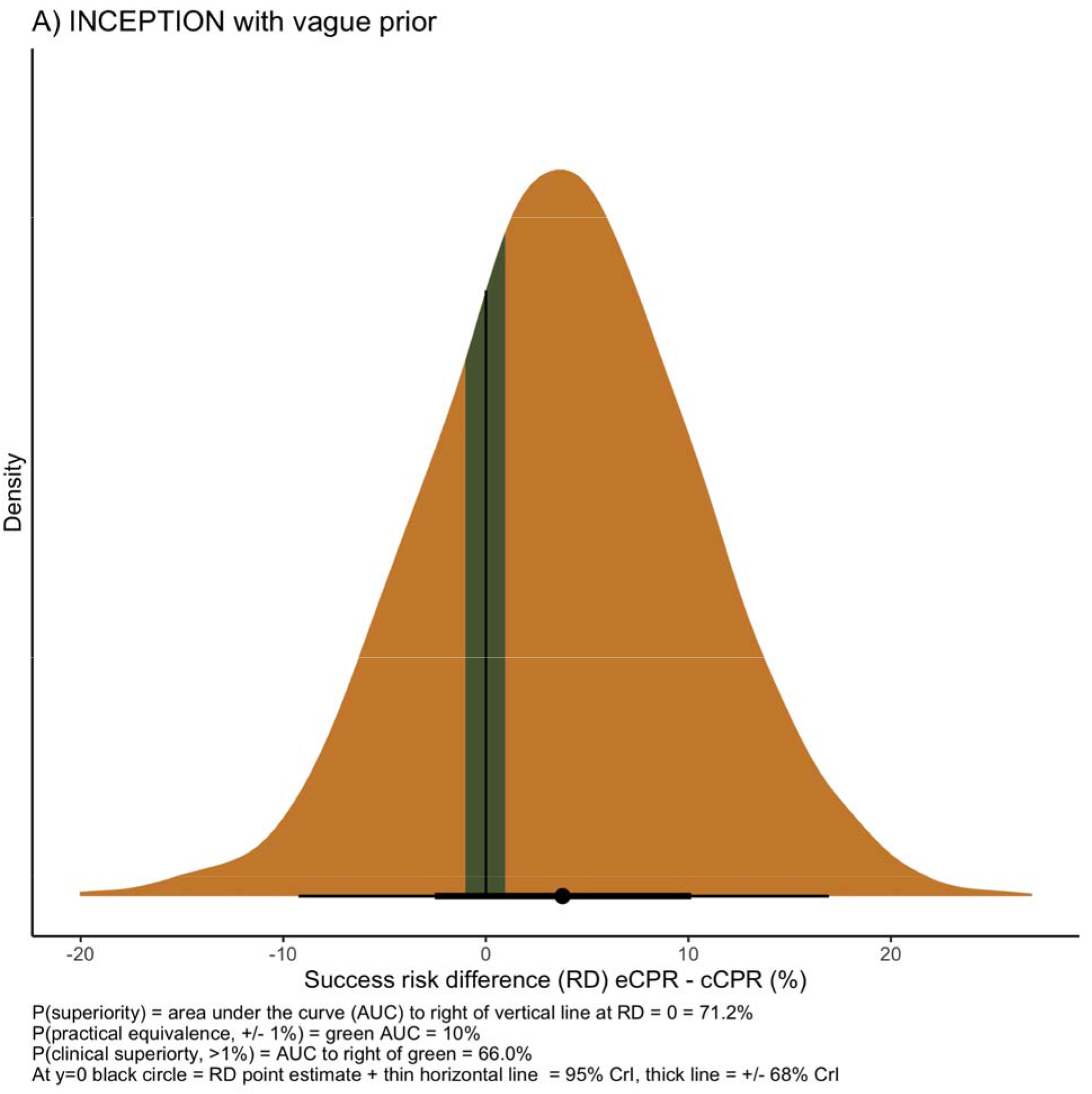
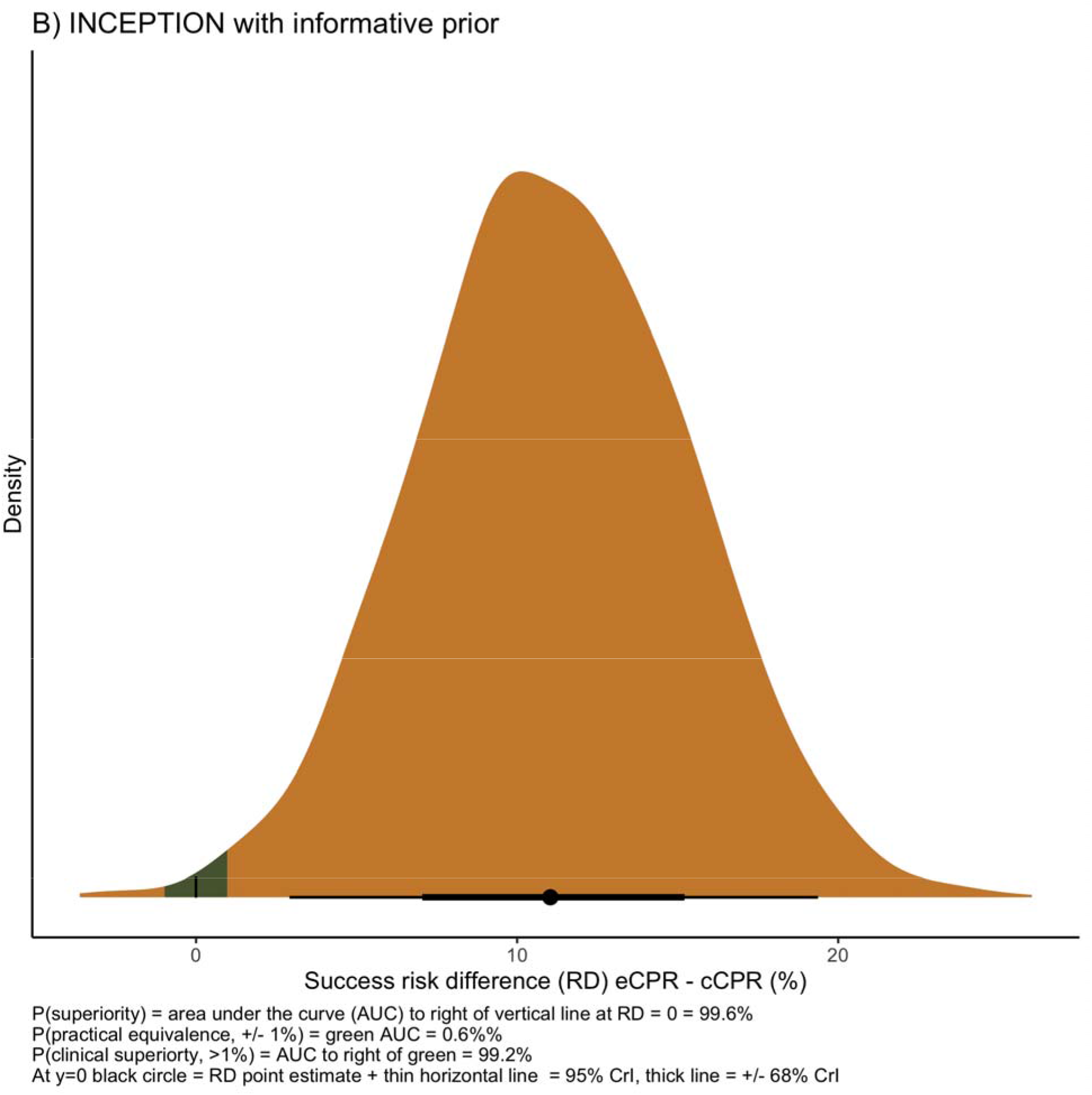
INCEPTION probability density functions (ITT analyses)

The ARREST(Yannopoulos et al. 2020) and PRAGUE(Belohlavek et al. 2022) trials have also randomized out of hospital cardiac arrest patients to cCPR to eCPR and their data may constitute informed priors for an updated Bayesian analysis of the INCEPTION(Suverein et al. 2023) data. This combined prior data can be summarized as a beta distribution with 25 successes and 122 failures for the cCPR arm and 44 successes and 94 failures for the eCPR arm. As a sensitivity analysis we performed a Bayesian analysis using as prior information each trial individually. The ARREST(Yannopoulos et al. 2020) data may be summarized as a beta distribution with 1 successes and 14 failures for the cCPR arm and 6 successes and 8 failures for the eCPR arm and considered as an enthusiastic prior, since this trial was halted prematurely for efficacy. Similarly the PRAGUE(Belohlavek et al. 2022) data can be summarized as a beta distribution with 24 successes and 108 failures for the cCPR arm and 38 successes and 86 failures for the eCPR arm and considered as an skeptical prior, since this trial was halted prematurely for futility based on a lack of benefit at 6 months. The posterior distributions for these different priors are also shown in Table 1 and ranged from 93.5% to 99.6% depending on the informative prior chosen (Table 1 and Figure 1B). Similarly under all scenarios with informed priors, the probability that the eCPR benefit exceeded the clinical threshold of a minimal 1% RD improvement exceeded 90% and the range of equivalence was consequentially always < 10%. Regardless of the informed prior chosen, the inclusion of additional data leads to a reduction in the uncertainty with narrower posterior 95% CrIs compared to the what was observed with the vague prior.

With the INCEPTION(Suverein et al. 2023) per-protocol data and a vague prior, the RD of increased survival with eCPR compared to cCPR is reversed (point estimate = -13.7) but with very wide 95% CrIs (−37.8 to 10.5. With the combined informative prior, per protocol analyses resulted in reduced posterior probabilities for eCPR benefit (86.4%) and an increased probability of equivalence (8.2%) due to the markedly different and decreased eCPR INCEPTION(Suverein et al. 2023) success rates thereby limiting definitive conclusions (full per protocol results available at https://github.com/brophyj/eCPR).

## Discussion

This Bayesian analysis of the INCEPTION(Suverein et al. 2023) data with a vague prior, shows a non-negligible probability of a potentially clinically meaningful mortality benefit with eCPR compared to cCPR. When combined with pertinent prior information the posterior probability for improved eCPR 30 day survival is 93-99%. Depending on the chosen prior, the posterior probability of at least a 1% absolute RD improvement in 30 day survival with eCPRis iexceeds 90%. This analysis suggests only a small probability that differences between the two techniques are within a reasonably acceptable equivalency range. Using only the INCEPTION per protocol data and a vague prior shows only a trivially small probability for a eCPR benefits. The inclusion of an informative priors suggests a moderate 86% probability a clinicially meaninngful eCPR benefit.

The INCEPTION(Suverein et al. 2023) researchers addressed an important and challenging clinical question and are to be congratulated on their trial design, execution, and nuanced discussion. However the constraints of standard statistical analyses limit a quantitative appreciation of the data, prevent an updating of past knowledge and encourage the perception that trials that fail to meet statistical significance are “negative” trials.

In contrast, Bayesian analyses concentrate on direct estimation of the parameters of interest accompanied by direct probability statements regarding their uncertainty (herein the risk of 30 day survival with intact neurological status), as well as permitting the incorporation prior knowledge (Brophy 2021; Zampieri et al. 2021). The avoidance of dichotomization minimizes the confusion “between absence of evidence and evidence of absence”(Altman and Bland 1995).

This Bayesian analysis has limitations. We did not have access to individual data thereby limiting any subgroup analyses. Moreover while the different RCTs in theory examined the same interventions, there are obvioulsy nuanced differences in their applications and the populations studied. Also the threshold choices for superiority and equivalence are arbitrary, although the analyses can be easily repeated for different choices. We choose not to perform a traditional Bayesian meta-analysis with a semi-informative prior for the between-study variation because our goal was to illustrate how Bayesian principles could be informative when applied to the analysis and interpretation of new trial data, in situations both with and without previous knowledge.

This analysis suggests a conclusion of “similar survival effects of eCPR to cCPR”(Suverein et al. 2023) is inaccurate and rather there is a high probability of a clinically meaningful 30 day benefit with eCPR, albeit with a slightly reduced per protocol probability benefit. However as shown by the reversal at 6 month of the encouraing 30 day PRAGUE data, the probability of persistent clinically meaningful longer term eCPR benefits remain to be demonstrated. The final dilemma confronting clinicians is that even Bayesian analyses provide only probability estimates for average treatment effects and not for the more elusive individual treatment effect.

## Data Availability

all data is secondary data that has been previously published

https://github.com/brophyj/eCPR

## Supplemental material

**Table 2.**
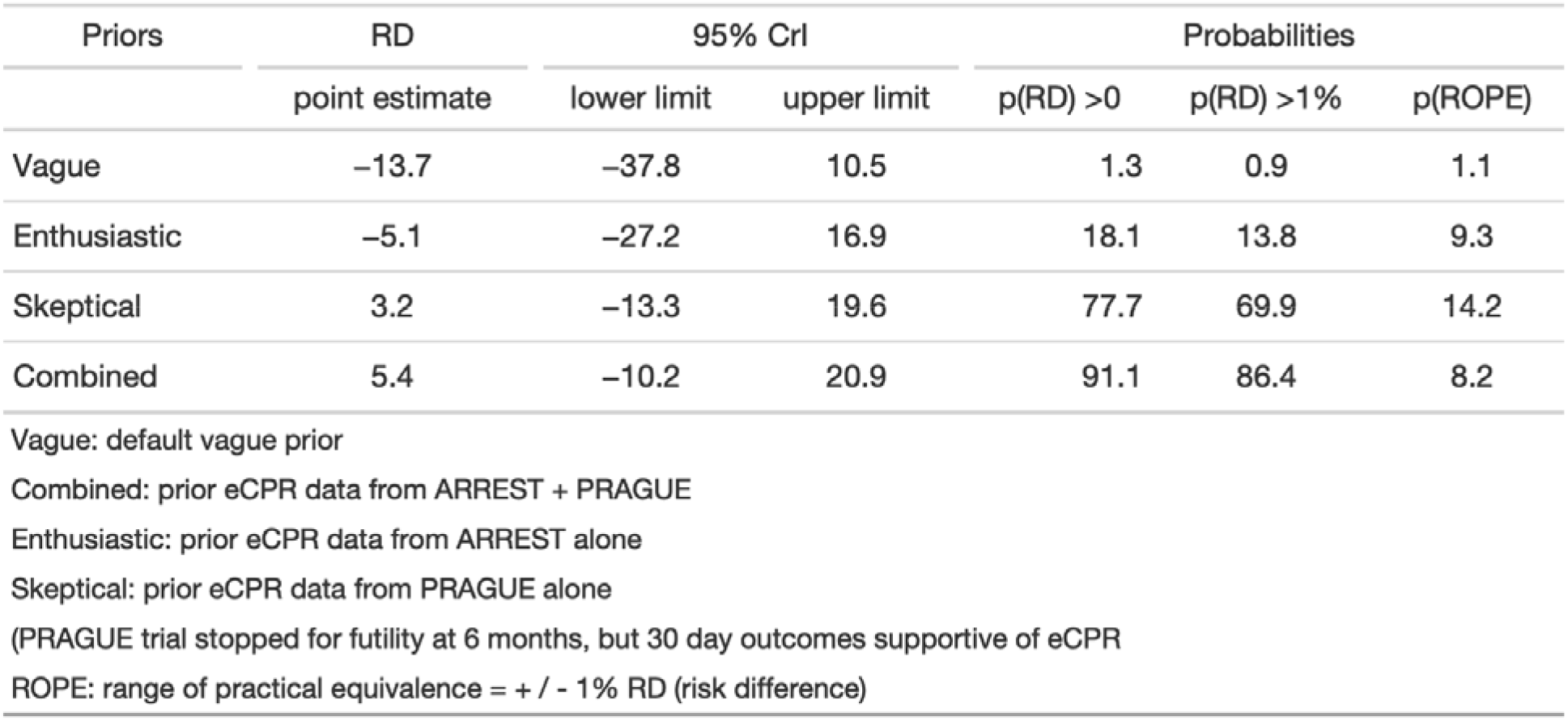
eCPR risk differences 95% credible intervals and probabilities with various priors per protocol analyses.

| Priors | RD<br>point estimate | 95% CrI |  | Probabilities |  |  |
| --- | --- | --- | --- | --- | --- | --- |
|  |  | lower limit | upper limit | p(RD) >0 | p(RD) >1% | p(ROPE) |
| Vague | -13.7 | -37.8 | 10.5 | 1.3 | 0.9 | 1.1 |
| Enthusiastic | -5.1 | -27.2 | 16.9 | 18.1 | 13.8 | 9.3 |
| Skeptical | 3.2 | -13.3 | 19.6 | 77.7 | 69.9 | 14.2 |
| Combined | 5.4 | -10.2 | 20.9 | 91.1 | 86.4 | 8.2 |

Vague: default vague prior
Combined: prior eCPR data from ARREST + PRAGUE
Enthusiastic: prior eCPR data from ARREST alone
Skeptical: prior eCPR data from PRAGUE alone
(PRAGUE trial stopped for futility at 6 months, but 30 day outcomes supportive of eCPR)
ROPE: range of practical equivalence = + / - 1% RD (risk difference)

**Figure 2.**
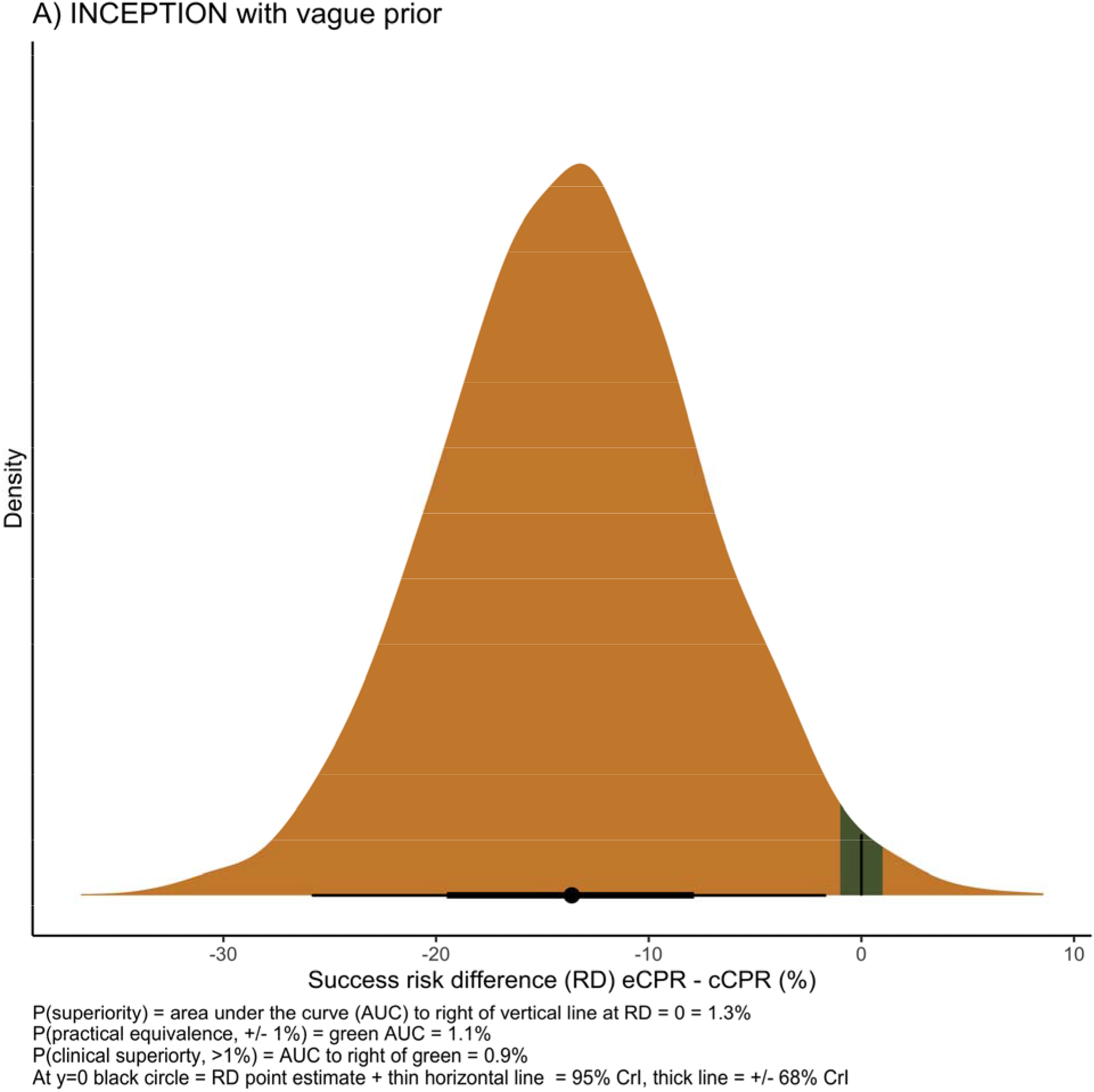
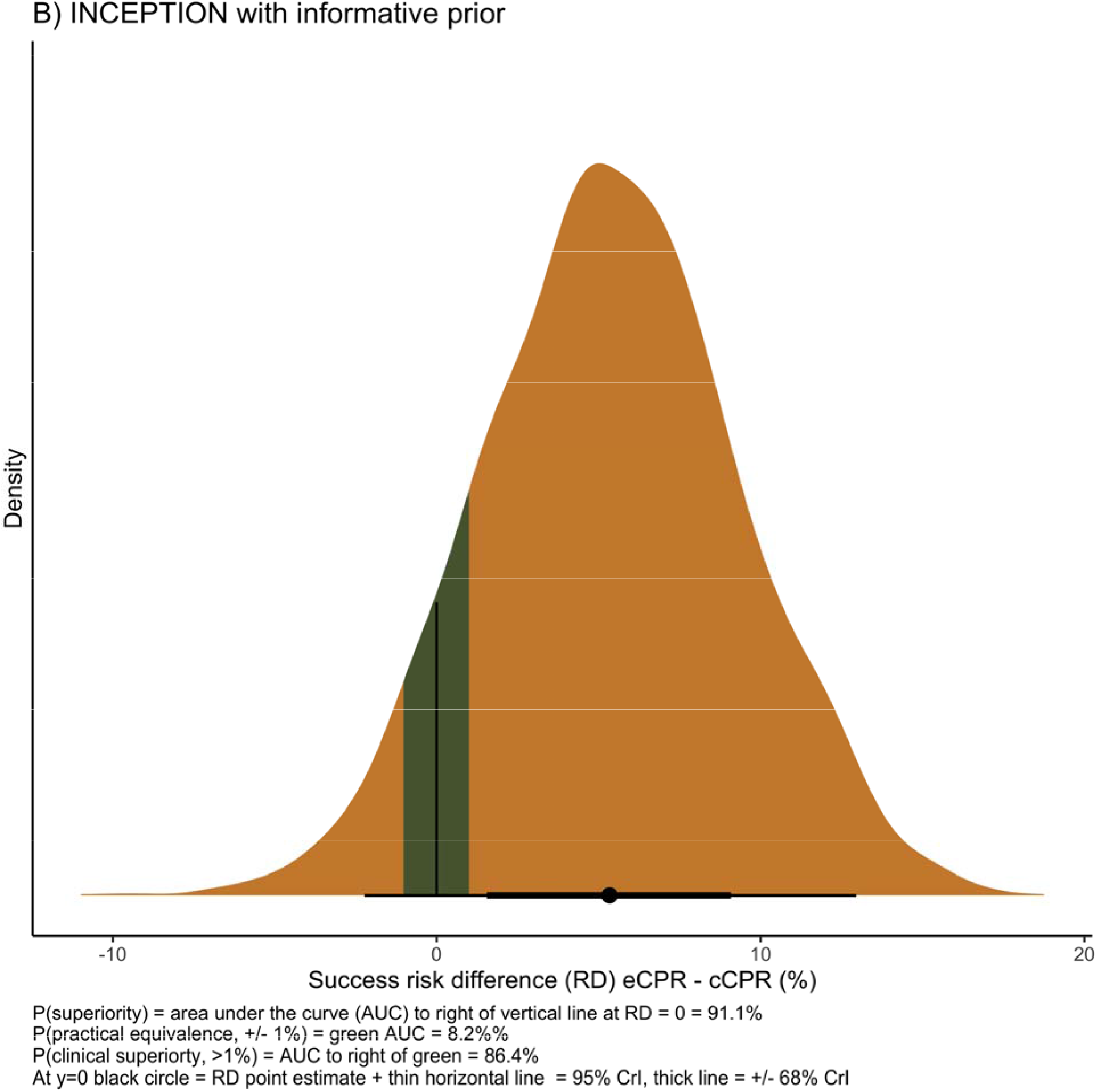
INCEPTION probability density functions (PP analyses)

